# Multidimensional Sleep Profiles via Machine learning and Risk of Dementia and Cardiovascular Disease

**DOI:** 10.1101/2024.08.19.24312248

**Authors:** Clémence Cavaillès, Meredith Wallace, Yue Leng, Katie L. Stone, Sonia Ancoli-Israel, Kristine Yaffe

## Abstract

**Importance:** Sleep health comprises several dimensions such as duration and fragmentation of sleep, circadian activity, and daytime behavior. Yet, most research has focused on individual sleep characteristics. Studies are needed to identify sleep profiles incorporating multiple dimensions and to assess how different profiles may be linked to adverse health outcomes.

**Objective:** To identify actigraphy-based 24-hour sleep/circadian profiles in older men and to investigate whether these profiles are associated with the incidence of dementia and cardiovascular disease (CVD) events over 12 years.

**Design:** Data came from a prospective sleep study with participants recruited between 2003-2005 and followed until 2015-2016.

**Setting:** Multicenter population-based cohort study.

**Participants:** Among the 3,135 men enrolled, we excluded 331 men with missing or invalid actigraphy data and 137 with significant cognitive impairment at baseline, leading to a sample of 2,667 participants.

**Exposures:** Leveraging 20 actigraphy-derived sleep and circadian activity rhythm variables, we determined sleep/circadian profiles using an unsupervised machine learning technique based on multiple coalesced generalized hyperbolic mixture modeling.

**Main Outcomes and Measures:** Incidence of dementia and CVD events.

**Results:** We identified three distinct sleep/circadian profiles: active healthy sleepers (AHS; n=1,707 (64.0%); characterized by normal sleep duration, higher sleep quality, stronger circadian rhythmicity, and higher activity during wake periods), fragmented poor sleepers (FPS; n=376 (14.1%); lower sleep quality, higher sleep fragmentation, shorter sleep duration, and weaker circadian rhythmicity), and long and frequent nappers (LFN; n=584 (21.9%); longer and more frequent naps, higher sleep quality, normal sleep duration, and more fragmented circadian rhythmicity). Over the 12-year follow-up, compared to AHS, FPS had increased risks of dementia and CVD events (Hazard Ratio (HR)=1.35, 95% confidence interval (CI)=1.02-1.78 and HR=1.32, 95% CI=1.08-1.60, respectively) after multivariable adjustment, whereas LFN showed a marginal association with increased CVD events risk (HR=1.16, 95% CI=0.98-1.37) but not with dementia (HR=1.09, 95%CI=0.86-1.38).

**Conclusion and Relevance:** We identified three distinct multidimensional profiles of sleep health. Compared to healthy sleepers, older men with overall poor sleep and circadian activity rhythms exhibited worse incident cognitive and cardiovascular health. These results highlight potential targets for sleep interventions and the need for more comprehensive screening of poor sleepers for adverse outcomes.

**Key Points:**

- Question: Are there distinct sleep/circadian profiles in older men, and if so, are they associated with the incidence of dementia and cardiovascular disease (CVD) events over 12 years?
- Findings: Three actigraphy-based profiles were identified: active healthy sleepers [AHS], fragmented poor sleepers [FPS], and long and frequent nappers [LFN]. Compared to AHS, FPS had increased risks of dementia and CVD events whereas LFN had marginal risk of CVD events.
- Meaning: Older men with distinct sleep/circadian profiles are at increased risk of incident dementia and CVD events, suggesting their potential as target populations for sleep interventions and screening for adverse outcomes.

## Introduction

Growing evidence has linked individual sleep characteristics and disturbed circadian rhythms with adverse health outcomes in older adults, including neurodegenerative and cardiovascular diseases (CVDs), two leading causes of disability and mortality worldwide.^1–4^ However, the literature remains inconsistent.^5–8^ Some studies have associated both short and long sleep duration with increased dementia risk,^9^ while others found conflicting associations.^7,10,11^ Similarly, although some research has suggested that more frequent or long naps were associated with a higher risk of CVD,^4,12^ others showed a protective effect.^13^

These conflicting findings may be partly due to the lack of consideration of the multidimensional nature of sleep. Research has primarily examined sleep characteristics in isolation, whereas sleep involves a complex interplay of multiple dimensions such as duration, continuity, quality, circadian rhythmicity, and napping.^14^ Adopting a holistic approach by considering common combinations of sleep characteristics could improve our understanding of multidimensional sleep patterns and their associations with outcomes. Investigating these associations can provide valuable insights for public health strategies, aiding the identification of at-risk populations and targeted treatments or interventions.

In a community-dwelling cohort of older men, our objectives were: (1) to identify actigraphy-derived sleep health profiles based on multidimensional objective sleep and rest-activity variables, by using a novel and flexible clustering method; and (2) to investigate the longitudinal associations between these profiles and the incidence of dementia and CVD events over 12 years.

## Methods

We followed the Strengthening the Reporting of Observational Studies in Epidemiology (STROBE) reporting guidelines.

### Study Design

From 2000 to 2002, the Osteoporotic Fractures in Men Study (MrOS) enrolled 5,994 community-dwelling men aged ≥65 years, able to walk without assistance, and without bilateral hip replacements, at six clinical centers across the United States.^15,16^ Among them, 3,135 were recruited into the ancillary MrOS sleep study and underwent a comprehensive sleep assessment between 2003 and 2005 (our study baseline). We excluded 331 men with missing or invalid actigraphy data (< 3 “in-bed” and “out-of-bed” intervals), and 137 with significant cognitive impairment at baseline (Modified Mini-Mental State Examination (3MS) score <80 or taking dementia medication), leading to a sample of 2,667 participants (eFigure 1). All participants provided written informed consent and the study was approved by the Institutional Review Board at each site.

### Actigraphy

Participants wore a SleepWatch-O actigraph (Ambulatory Monitoring, Inc) continuously on their nondominant wrist for ≥4 consecutive 24-hours periods (median 5, range 3-13). Data were collected in proportional integration mode and scored by epoch to estimate “wake” and “sleep” periods using Action W-2 software and the University of California, San Diego scoring algorithm.^17^ Trained scorers at the San Francisco Coordinating Center edited the data using participants’ sleep diaries to identify time in and out of bed as well as periods when the interval should be deleted because the watch was removed. Sleep indices were summarized across the monitoring period using means and standard deviations (SDs).^18,19^ Circadian rest-activity rhythm indices were generated using parametric extended cosine models and nonparametric variables.^20,21^ A total of 37 actigraphy variables were examined and described in Table 1.

**Table 1.**
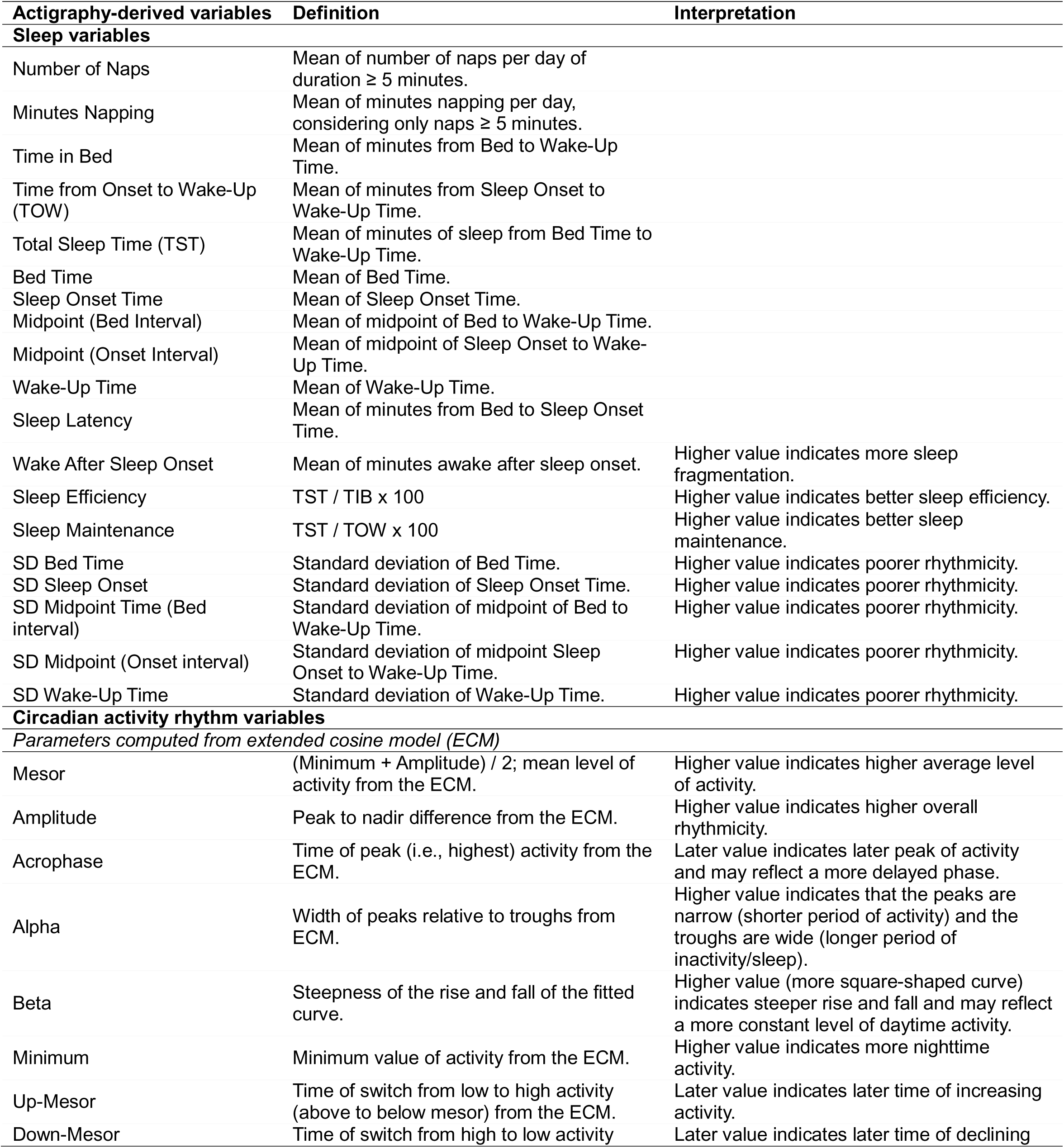
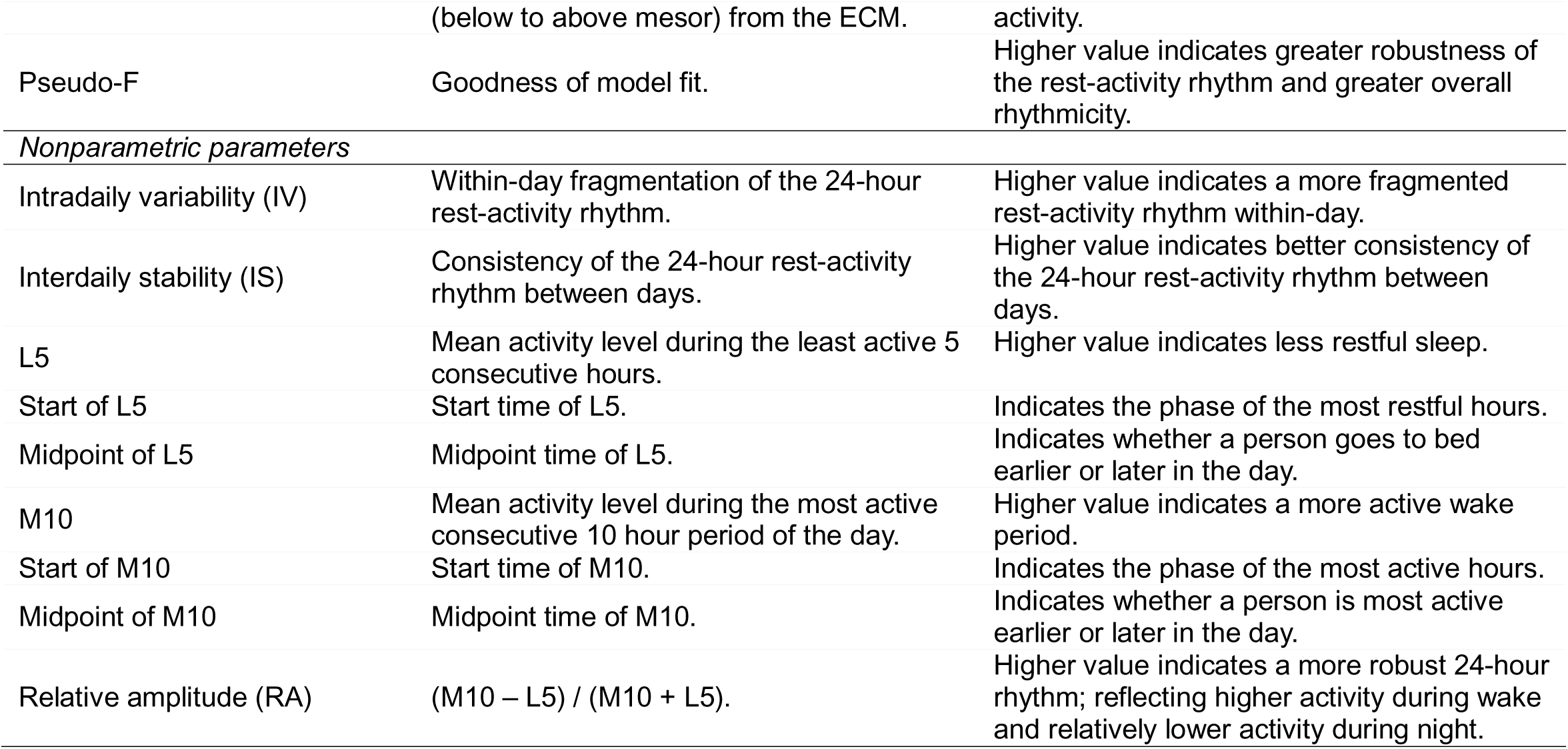
Description and interpretation of all actigraphy-derived variables.

### Dementia Incidence

Over 12 years, participants attended four follow-up visits where they reported any physician-diagnosed dementia and their medication use, bringing all medications taken within the past 30 days. Dementia medication use was categorized based on the Iowa Drug Information Service Drug Vocabulary.^22^ In addition, trained staff administered the 3MS test to assess global cognitive function. Incident dementia at any follow-up visit was defined by meeting at least one of the following criteria: (i) self-reported physician-diagnosed dementia; (ii) dementia medication use; or (iii) a change in 3MS score of ≥1.5 SDs worse than the mean change from baseline to any follow-up visit. Participants were censored at the date of the diagnostic visit, death, or last visit.

### Cardiovascular Disease Event Incidence

Participants were surveyed for incident CVD events by postcard and phone contact every four months for approximately 12 years, with a response rate over 99%. Relevant medical records and documentation from any potential incident clinical events were obtained by the clinical center. For both nonfatal and fatal CVD events, all documents were adjudicated by a board-certified cardiologist using a prespecified adjudication protocol. Inter-rater agreement was periodically evaluated by one or more expert adjudicator(s) in a random subset of events to ensure quality control. Confirmed incident all-cause CVD events combined coronary heart disease, cerebrovascular, and peripheral vascular disease events (eMethod). Participants were censored at the date of the first CVD event, death, last contact before March 1, 2015, or on March 1, 2015.

### Statistical Analysis

We conducted a cluster analysis to identify distinct sleep/circadian profiles. Firstly, we selected 20 actigraphy variables, choosing one of the two variables when their correlation was above 0.70 (eFigure 2). Secondly, we performed a principal component analysis (PCA) on the selected variables to reduce data dimensionality (while preserving most of the data variation) and enhance the efficacy of subsequent clustering. The number of principal components was determined considering components with eigenvalues >1 and by visual inspection of the scree plot (eFigure 3).^23,24^ Thirdly, sleep/circadian profiles were identified using Multiple Coalesced Generalized Hyperbolic Distribution (MixGHD package in R) mixture models.^25,26^ This method, as opposed to standard clustering approaches, was chosen for its ability to accommodate potentially skewed and/or asymmetric clusters, an important consideration given the skewed distributions often observed in actigraphy data (eFigure 4). We explored models comprising one to five clusters, using k-medoids as the starting criterion, and determined the optimal number of clusters by examining the Bayesian Information Criteria (BIC), the Akaike Information Criteria (AIC) and the Integrated Complete-data Likelihood (ICL) (see eMethods for additional information).

We performed unadjusted and multivariable adjusted Cox proportional hazards models with age as time scale to investigate whether identified sleep profiles were associated with the incidence of dementia and CVD events over 12 years. Covariates were selected based on potential biological plausibility, and included study site, race/ethnicity, education, smoking status, caffeine intake, alcohol use, physical activity, body mass index (BMI), history of diabetes mellitus and hypertension, depressive symptoms, and sleep-related medications use (eMethods).

In sensitivity analyses, models were further adjusted for (i) history of heart attack and stroke, (ii) baseline apnea-hypopnea index (AHI), and (iii) baseline 3MS score (for dementia analysis). We also excluded participants with incident dementia at the first follow-up visit to minimize reverse causation (for dementia analysis), and those with history of heart attack and stroke to minimize confounding bias (for CVD analysis).

Significance level was set at a two-sided *p* < 0.05 and statistical analyses were performed using R version 4.3.0.

## Results

A total of 2,667 men were eligible for cluster analysis. At baseline, participants had a median age of 75 years (interquartile range [IQR]= 72-80), 20.2% had a high school education or lower, and 90.0% were White. Compared to included participants, excluded men (n=468) were older, less educated, more likely to be non-White, and had less physical activity and alcohol consumption, but higher depressive symptoms and sleep medication use (eTable 1).

### Sleep profiles

After examining the AIC, BIC, and ICL, three distinct sleep/circadian profiles were identified (eFigure 5): active healthy sleepers [AHS; n=1,707 (64.0%)], fragmented poor sleepers [FPS; n=376 (14.1%)], and long and frequent nappers [LFN; n=584 (21.9%)]. All sleep characteristics are described in Table 2.

**Table 2.** Sleep characteristics among the 2,667 participants according to identified multidimensional sleep clusters.

|  | Active Healthy Sleepers<br>(n=1707) | Fragmented Poor Sleepers<br>(n=376) | Long and Frequent Nappers<br>(n=584) |  |  |  |
| --- | --- | --- | --- | --- | --- | --- |
| Sleep variables | Median (IQR) | Median (IQR) | Median (IQR) | Effect size ($\eta^2$) | p-value<sup>c</sup> | Post hoc<sup>d</sup> |
| **Variables used to create the clusters** |  |  |  |  |  |  |
| Minutes Napping | 34 (16;60) | 31 (13;53) | 79 (48;127) | 0.160 | <.0001 | LFN > AHS,FPS |
| Time in Bed | 489 (453;524) | 530 (482;570) | 475 (440;514) | 0.056 | <.0001 | FPS > AHS > LFN |
| Total Sleep Duration | 400 (362;438) | 336 (273;395) | 381 (342;424) | 0.074 | <.0001 | AHS > LFN > FPS |
| Sleep Onset Time | 23.1 (22.4;23.7) | 23.7 (22.7;24.9) | 23.4 (22.8;24.1) | 0.037 | <.0001 | FPS > LFN > AHS |
| Wake-Up Time | 6.9 (6.2;7.5) | 7.5 (6.6;8.1) | 6.9 (6.4;7.5) | 0.028 | <.0001 | FPS > AHS,LFN |
| SD Sleep Onset | 0.57 (0.37;0.83) | 1.14 (0.80;1.64) | 0.58 (0.37;0.90) | 0.131 | <.0001 | FPS > AHS,LFN |
| SD Wake-Up Time | 0.56 (0.37;0.80) | 0.72 (0.44;1.16) | 0.49 (0.31;0.77) | 0.027 | <.0001 | FPS > AHS > LFN |
| Sleep Latency | 18 (11;30) | 53 (31;96) | 19 (11;32) | 0.157 | <.0001 | FPS > AHS,LFN |
| Wake After Sleep Onset | 61 (42;85) | 126 (97;159) | 65 (43;93) | 0.188 | <.0001 | FPS > AHS,LFN |
| Mesor<sup>a</sup> | 2102 (1850;2359) | 2170 (1863;2433) | 2257 (1893;2678) | 0.020 | <.0001 | LFN > FPS > AHS |
| Pseudo-F<sup>a</sup> | 1078 (781;1421) | 861 (637;1169) | 805 (558;1109) | 0.065 | <.0001 | AHS > FPS > LFN |
| Beta<sup>a</sup> | 8.03 (4.89;17.02) | 8.59 (5.02;20.14) | 12.33 (4.43;40.20) | 0.007 | <.0001 | LFN > AHS,FPS |
| Alpha<sup>a</sup> | -0.40 (-0.50;-0.28) | -0.35 (-0.46;-0.22) | -0.09 (-0.25;0.10) | 0.213 | <.0001 | LFN > FPS > AHS |
| Acrophase<sup>a</sup> | 14.38 (13.77;14.98) | 14.69 (13.89;15.57) | 13.51 (12.90;14.16) | 0.114 | <.0001 | FPS > AHS > LFN |
| Minimum<sup>a</sup> | 234 (3;385) | 521 (265;756) | 588 (330;781) | 0.193 | <.0001 | FPS,LFN > AHS |
| Intradaily variability (IV)<sup>b</sup> | 0.60 (0.48;0.72) | 0.63 (0.51;0.77) | 0.70 (0.54;0.85) | 0.037 | <.0001 | LFN > FPS > AHS |
| Interdaily stability (IS)<sup>b</sup> | 0.76 (0.69;0.82) | 0.72 (0.64;0.79) | 0.73 (0.66;0.80) | 0.021 | <.0001 | AHS > FPS,LFN |
| Start of M10<sup>b</sup> | 8.2 (7.3;9.2) | 8.7 (7.7;10.1) | 7.6 (6.9;8.6) | 0.039 | <.0001 | FPS > AHS > LFN |
| Start of L5<sup>b</sup> | 0.23 (-0.70;1.13) | 0.73 (-0.38;1.97) | 0.55 (-0.27;1.35) | 0.016 | <.0001 | FPS,LFN > AHS |
| Midpoint of L5<sup>b</sup> | 2.73 (1.80;3.63) | 3.23 (2.12;4.47) | 3.05 (2.23;3.85) | 0.016 | <.0001 | FPS,LFN > AHS |
| **Other variables** |  |  |  |  |  |  |
| Number of Naps | 2.5 (1.3;4.3) | 2.3 (1.1;3.8) | 5.5 (3.5;8.5) | 0.167 | <.0001 | LFN > AHS,FPS |
| Time from Onset to Wake-Up | 460 (423;495) | 453 (387;508) | 444 (409;484) | 0.009 | <.0001 | AHS > LFN |
| Bed Time | 22.7 (22.1;23.4) | 22.6 (21.8;23.5) | 23.0 (22.4;23.7) | 0.017 | <.0001 | LFN > AHS,FPS |
| SD Bed Time | 0.51 (0.33;0.75) | 0.68 (0.41;1.01) | 0.51 (0.31;0.83) | 0.020 | <.0001 | FPS > AHS,LFN |
| Midpoint (Bed Interval) | 2.79 (2.24;3.33) | 2.95 (2.30;3.73) | 2.97 (2.44;3.52) | 0.011 | <.0001 | FPS,LFN > AHS |
| Midpoint (Onset Interval) | 2.92 (2.37;3.48) | 3.48 (2.68;4.36) | 3.09 (2.57;3.65) | 0.038 | <.0001 | FPS > LFN > AHS |
| SD Midpoint (Bed interval) | 0.43 (0.30;0.61) | 0.58 (0.37;0.87) | 0.40 (0.25;0.63) | 0.026 | <.0001 | FPS > AHS,LFN |
| SD Midpoint (Onset interval) | 0.45 (0.30;0.63) | 0.75 (0.53;1.05) | 0.43 (0.27;0.68) | 0.089 | <.0001 | FPS > AHS,LFN |
| Sleep Efficiency | 83 (77;87) | 64 (54;72) | 82 (75;87) | 0.218 | <.0001 | AHS > LFN > FPS |
| Sleep Maintenance | 87 (82;91) | 71 (62;79) | 85 (79;90) | 0.190 | <.0001 | AHS > LFN > FPS |
| Amplitude<sup>a</sup> | 3712 (3154;4280) | 3294 (2611;3894) | 3250 (2591;4125) | 0.038 | <.0001 | AHS > FPS,LFN |
| Up-Mesor<sup>a</sup> | 6.9 (6.3;7.5) | 7.4 (6.6;8.1) | 7.0 (6.5;7.9) | 0.028 | <.0001 | FPS > LFN > AHS |
| Down-Mesor<sup>a</sup> | 22.0 (21.1;22.8) | 22.0 (21.0;23.1) | 19.9 (18.9;20.9) | 0.202 | <.0001 | AHS,FPS > LFN |
| M10<sup>b</sup> | 4049 (3543;4534) | 3884 (3370;4406) | 3736 (3111;4332) | 0.023 | <.0001 | AHS > FPS > LFN |
| Midpoint of M10<sup>b</sup> | 13.2 (12.3;14.2) | 13.7 (12.7;15.1) | 12.6 (11.9;13.6) | 0.039 | <.0001 | FPS > AHS > LFN |
| L5<sup>b</sup> | 292 (217;383) | 525 (414;682) | 323 (232;442) | 0.164 | <.0001 | FPS > LFN > AHS |
| Relative amplitude (RA)<sup>b</sup> | 0.86 (0.82;0.90) | 0.77 (0.68;0.81) | 0.84 (0.78;0.88) | 0.169 | <.0001 | AHS > LFN > FPS |

Abbreviations: IQR, interquartile range.
<sup>a</sup> computed from extended cosine model; <sup>b</sup> nonparametric measures; <sup>c</sup> Kruskal-Wallis test was used; <sup>d</sup> Dunn test adjusted for multiple comparisons using Bonferroni method was used.

AHS were characterized by normal nighttime sleep duration (median= 6.7 hours), higher sleep quality (median sleep efficiency= 83%, sleep maintenance= 87%, minimum= 234, L5= 292), earlier timing of sleep (median sleep onset time= 23.1, start and midpoint of L5= 0.23 and 2.73, midpoint of bed and onset interval= 2.79 and 2.92), stronger circadian rhythmicity (median amplitude= 3712, pseudo-F= 1078, intradaily variability=0.60, interdaily stability= 0.76, relative amplitude= 0.86), and higher activity during wake periods (median M10= 4049, alpha= −0.40) (see Table 1 for description and interpretation of sleep data).

FPS were characterized by shorter nighttime sleep duration (median= 5.6 hours) and longer time in bed (median= 8.8 hours), lower sleep quality (median sleep efficiency= 64%, sleep maintenance= 71), higher sleep fragmentation (median sleep latency= 53 min, wake after sleep onset= 126 min, L5= 525, and median SD for sleep onset= 1.14, bedtime= 0.68, wake-up time= 0.72, midpoint of bed and onset interval= 0.58 and 0.75), later timing of sleep and activity (median acrophase= 14.69, wake-up time= 7.5, start of M10= 8.7, up-mesor= 7.4, sleep onset time= 23.7, midpoint of onset interval= 3.48), and weaker circadian rhythmicity (median amplitude= 3294, relative amplitude= 0.77).

LFN were characterized by longer (median= 79 min) and more frequent naps (median= 5.5), normal nighttime sleep duration (median= 6.4 hours), good sleep quality (median sleep efficiency= 82%, sleep maintenance= 85%), earlier timing of activity (median acrophase= 13.51, start and midpoint of M10= 7.6 and 12.6, down-mesor= 19.9), and more fragmented circadian rhythmicity (median pseudo-F= 805, intradaily variability= 0.70, interdaily stability= 0.73, amplitude= 3250).

All sleep and circadian variables differed significantly across the three profiles (p<0.0001). Among the cluster analysis variables, large effect sizes were found for minutes napping, sleep latency, wake after sleep onset, alpha, and minimum; with the circadian variables alpha η^2^ =0.213) and minimum (η^2^ =0.193) being the largest contributors. Other variables with large effect sizes included number of naps, sleep efficiency, sleep maintenance, down-mesor, L5, and relative amplitude (Table 2). Sleep profiles based on the largest contributors were illustrated in Figure 1.

**Figure 1.**
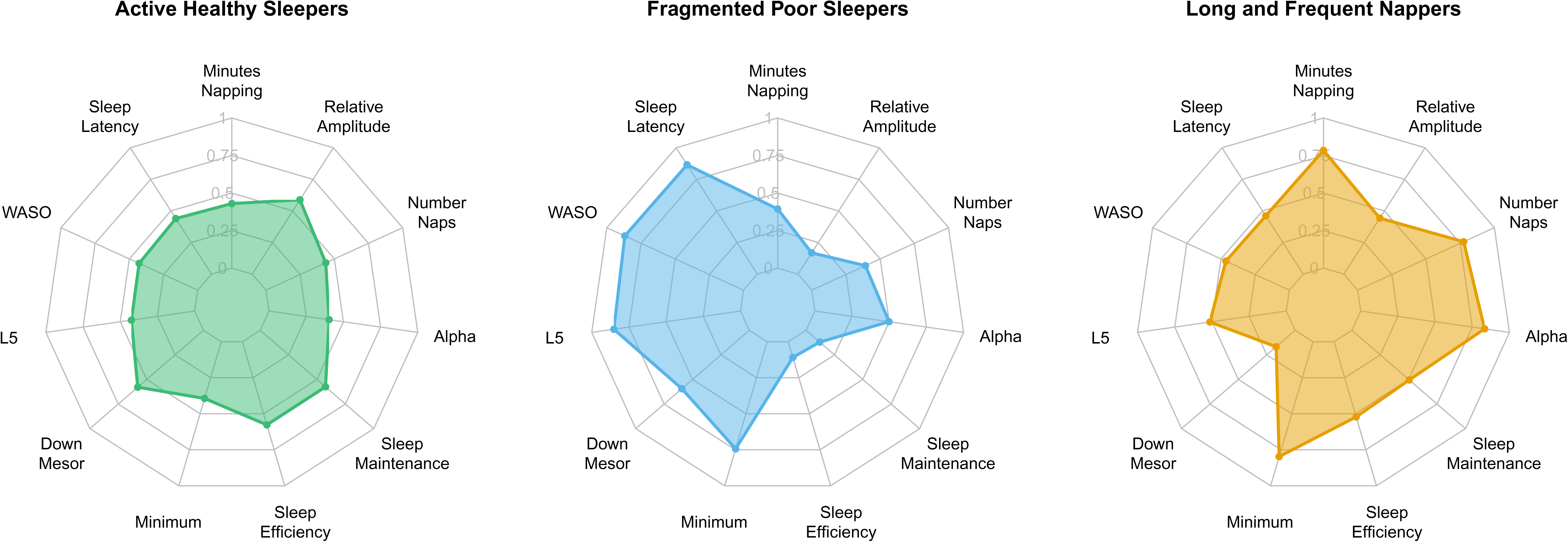
Radial plots displaying the median quantile rankings of sleep and circadian characteristics with large effect sizes for each sleep profile. The sample’s highest ranked value is represented by the maximum value of 1, the median ranked value by 0.50, and the lowest ranked value by 0.

Compared to AHS, FPS were less educated and less physically active, while LFN were slightly older. Both FPS and LFN were more likely to be non-White, smokers, to have a history of hypertension and a higher BMI. AHS consumed less caffeine than FPS (Table 3).

**Table 3.**
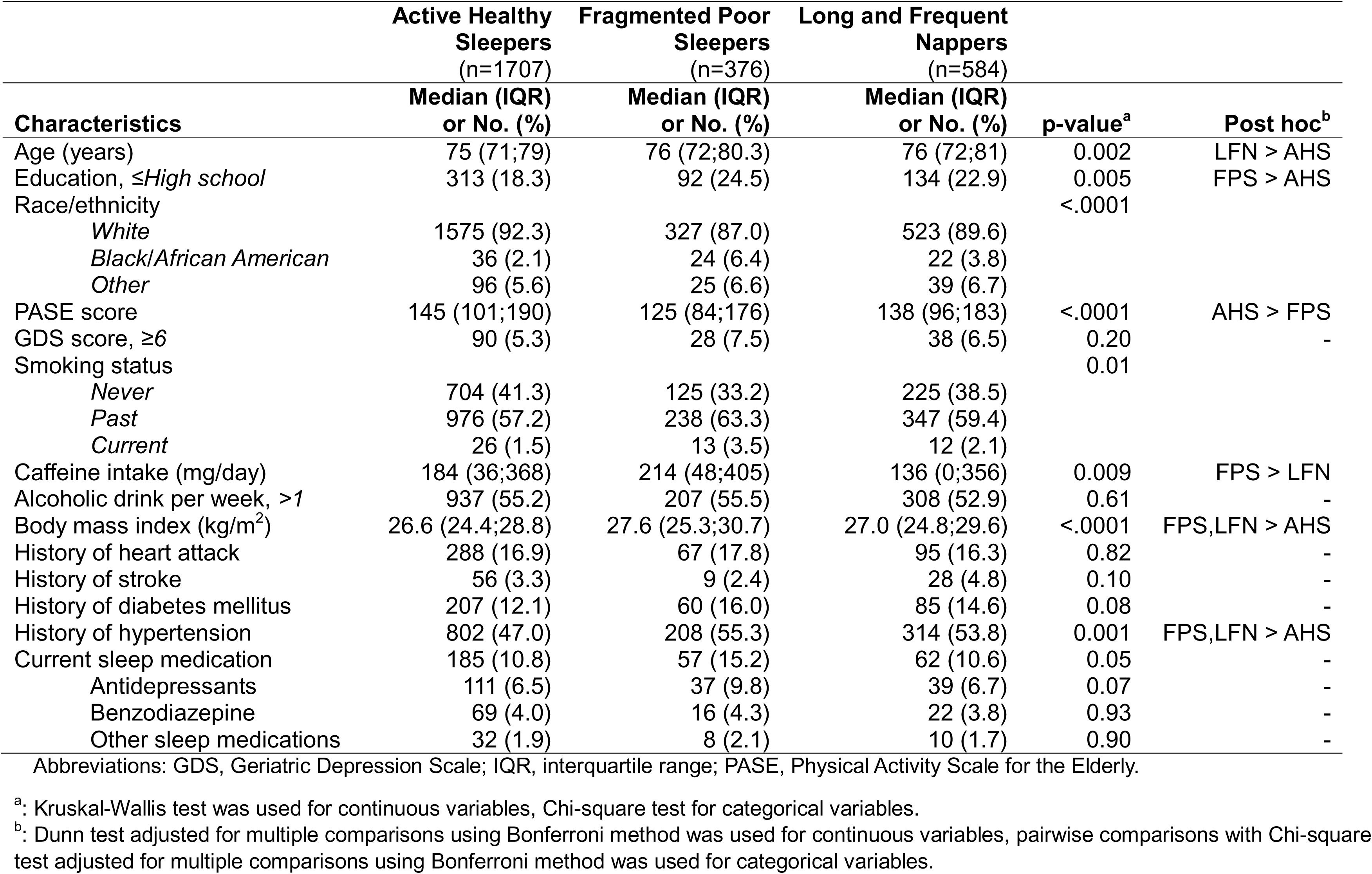
Baseline characteristics according to identified sleep clusters among the 2,667 participants.

|  | Active Healthy Sleepers<br>(n=1707) | Fragmented Poor Sleepers<br>(n=376) | Long and Frequent Nappers<br>(n=584) |  |  |
| --- | --- | --- | --- | --- | --- |
| Characteristics | Median (IQR)<br>or No. (%) | Median (IQR)<br>or No. (%) | Median (IQR)<br>or No. (%) | p-value<sup>a</sup> | Post hoc<sup>b</sup> |
| Age (years) | 75 (71;79) | 76 (72;80.3) | 76 (72;81) | 0.002 | LFN > AHS |
| Education, ≤High school | 313 (18.3) | 92 (24.5) | 134 (22.9) | 0.005 | FPS > AHS |
| Race/ethnicity |  |  |  | <.0001 |  |
| White | 1575 (92.3) | 327 (87.0) | 523 (89.6) |  |  |
| Black/African American | 36 (2.1) | 24 (6.4) | 22 (3.8) |  |  |
| Other | 96 (5.6) | 25 (6.6) | 39 (6.7) |  |  |
| PASE score | 145 (101;190) | 125 (84;176) | 138 (96;183) | <.0001 | AHS > FPS |
| GDS score, ≥6 | 90 (5.3) | 28 (7.5) | 38 (6.5) | 0.20 | - |
| Smoking status |  |  |  | 0.01 |  |
| Never | 704 (41.3) | 125 (33.2) | 225 (38.5) |  |  |
| Past | 976 (57.2) | 238 (63.3) | 347 (59.4) |  |  |
| Current | 26 (1.5) | 13 (3.5) | 12 (2.1) |  |  |
| Caffeine intake (mg/day) | 184 (36;368) | 214 (48;405) | 136 (0;356) | 0.009 | FPS > LFN |
| Alcoholic drink per week, $>1$ | 937 (55.2) | 207 (55.5) | 308 (52.9) | 0.61 | - |
| Body mass index (kg/m<sup>2</sup>) | 26.6 (24.4;28.8) | 27.6 (25.3;30.7) | 27.0 (24.8;29.6) | <.0001 | FPS, LFN > AHS |
| History of heart attack | 288 (16.9) | 67 (17.8) | 95 (16.3) | 0.82 | - |
| History of stroke | 56 (3.3) | 9 (2.4) | 28 (4.8) | 0.10 | - |
| History of diabetes mellitus | 207 (12.1) | 60 (16.0) | 85 (14.6) | 0.08 | - |
| History of hypertension | 802 (47.0) | 208 (55.3) | 314 (53.8) | 0.001 | FPS, LFN > AHS |
| Current sleep medication | 185 (10.8) | 57 (15.2) | 62 (10.6) | 0.05 | - |
| Antidepressants | 111 (6.5) | 37 (9.8) | 39 (6.7) | 0.07 | - |
| Benzodiazepine | 69 (4.0) | 16 (4.3) | 22 (3.8) | 0.93 | - |
| Other sleep medications | 32 (1.9) | 8 (2.1) | 10 (1.7) | 0.90 | - |

Abbreviations: GDS, Geriatric Depression Scale; IQR, interquartile range; PASE, Physical Activity Scale for the Elderly.
<sup>a</sup>: Kruskal-Wallis test was used for continuous variables, Chi-square test for categorical variables.
<sup>b</sup>: Dunn test adjusted for multiple comparisons using Bonferroni method was used for continuous variables, pairwise comparisons with Chi-square test adjusted for multiple comparisons using Bonferroni method was used for categorical variables.

### Dementia incidence

Among the 2,562 men with dementia data, 461 (18.0%) incident dementia cases were identified over 12 years of follow-up (median=6.1 [IQR= 3.2-10.5]). Kaplan-Meier curves are shown in Figure 2. In unadjusted models, FPS had an increased risk of dementia (hazard ratios (HR)=1.34, 95% confidence intervals (CI)=1.03-1.74) compared to AHS. There was no association with dementia risk for LFN (HR=1.11, 95% CI=0.89-1.39). After adjusting for demographics, behaviors, comorbidities and sleep medication use, results were similar (HR=1.35, 95% CI=1.02-1.78 for FPS and HR= 1.09, 95% CI=0.86-1.38 for LFN). Sensitivity analyses displayed comparable findings (eTable 2, 3, 4, and 5).

**Figure 2.**
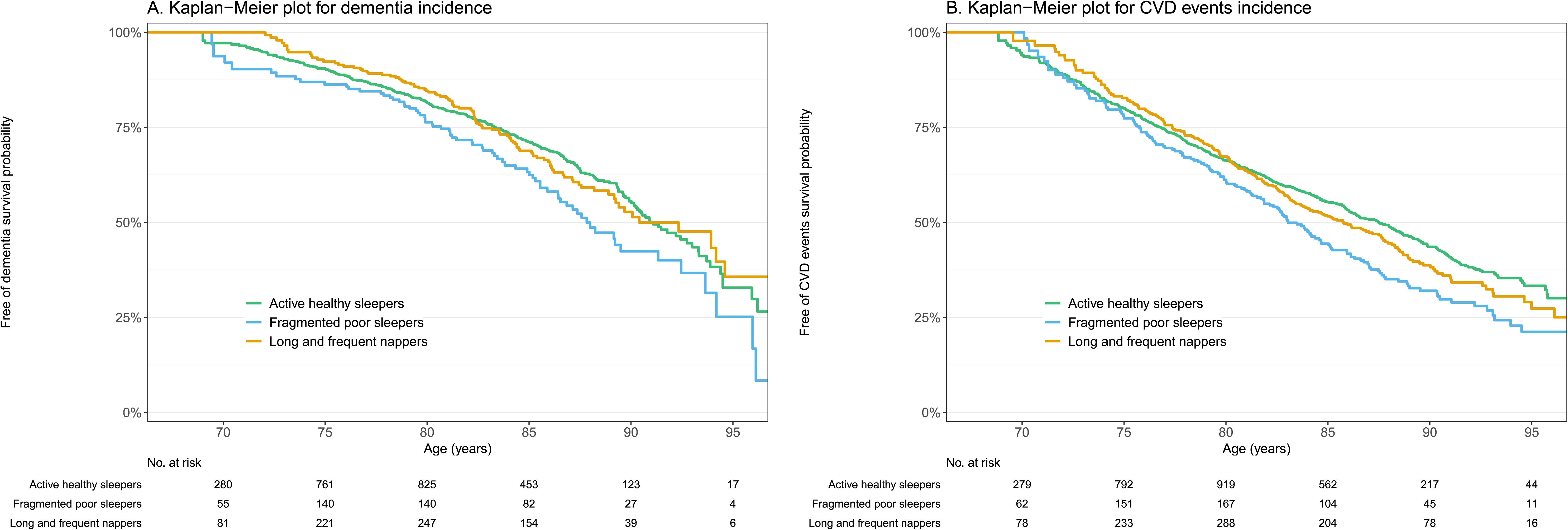
Kaplan-Meier curves depicting the probability of dementia-free and cardiovascular disease-free survival between sleep profiles.

### CVD event incidence

Among 2,606 men with CVD data, 839 (32.2%) incident CVD events were identified over 12 years of follow-up (median=9.7 [IQR= 4.5-10.5]). Kaplan-Meier curves are shown in Figure 2. In unadjusted models, both FPS and LFN were significantly associated with a higher risk of CVD events compared to AHS (HR=1.44, 95% CI=1.19-1.74 and HR=1.21, 95% CI=1.02-1.42, respectively). After multivariable adjustment, FPS were significantly associated with a higher risk of CVD events compared to AHS (HR=1.32, 95% CI=1.08-1.60), while LFN showed a borderline association (HR=1.16, 95% CI=0.98-1.37, p=0.08). Results remained consistent in the sensitivity analysis (eTable 2, 3 and 6), although the association for LFN was strongly attenuated after exclusion participants with a history of heart attack or stroke (eTable 6).

## Discussion

In a prospective cohort of older men, we identified three distinct multidimensional sleep/circadian profiles using machine learning: active healthy sleepers [AHS], fragmented poor sleepers [FPS], and long and frequent nappers [LFN]. Compared to AHS, FPS had increased risks of developing dementia and CVD events over 12 years whereas LFN tended to have an increased risk of CVD events, but not dementia. These results suggest that poor sleep and disrupted circadian rhythms may be risk factors or preclinical markers of dementia and CVD and highlight potential target populations for sleep interventions.

Few studies have used clustering^27–29^ or latent class^30–32^ analyses to discern sleep profiles in older adults. Moreover, these studies faced significant limitations, including cross-sectional design,^27^ reliance on self-reported sleep data,^27,30,31^ lack of rest-activity variables,^27,29–31^ and a focus on clinical populations.^29^ To the best of our knowledge, this study is the first to identify objective sleep and circadian profiles in community-dwelling older men using both sleep and rest-activity parameters with prospective follow-up for health outcomes. We identified three sleep profiles with high heterogeneity. The AHS group was the most common profile (64%), characterized by a combination of favorable characteristics: normal nighttime sleep duration, higher sleep quality, and stronger circadian rhythmicity. The LFN (21.9%) were characterized by longer and more frequent naps, alongside a combination of favorable and unfavorable dimensions: normal nighttime sleep duration, good sleep quality, and more fragmented circadian rhythms. The third group was the FPS (14.1%) who had a combination of unfavorable characteristics: shorter nighttime sleep duration, lower sleep quality, higher sleep fragmentation, delayed sleep/activity timing, and weaker circadian rhythmicity. Compared to prior research, our study provides a deeper characterization of nighttime and daytime sleep patterns by using a broader set of objective parameters, including extensive analysis of circadian rhythms. This provided a more nuanced and complete understanding of participants’ multidimensional sleep and circadian patterns. Additionally, the advanced machine learning technique has further enhanced classification accuracy.

Compared to AHS, FPS had a higher risk of dementia, consistent with variable-centered research linking short sleep duration, sleep fragmentation, poor sleep efficiency, and weak circadian rhythms with dementia incidence.^2,6,33–35^ This result is also in line with our previous work demonstrating the association between a multidimensional measure of sleep health and long-term cognitive decline.^36^ Our result extends those of a recent cross-sectional, person-centered study that used self-reported sleep, which found that the “poor sleepers” group performed worse on several cognitive tests compared to the “healthy sleepers” group.^27^ Potential underlying mechanisms include accumulation of amyloid-beta and tau proteins, disturbed glymphatic clearance, metabolic dysfunction, and inflammation.^37–39^ However, we cannot exclude the fact that preclinical dementia-related changes may also influence sleep and circadian patterns.^40–42^ FPS also had an increased risk of CVD events, in line with several prior studies of individual sleep parameters.^43–45^ Increase sympathetic activity and blood pressure, disrupted endothelial function, and inflammatory processes may explain in part this association.^46^ Taken together, these results showed that FPS were associated with poor incident cognitive and cardiovascular health.

We did not observe an association between LFN and dementia incidence. This finding contributes to the ongoing debate on napping and dementia. Some studies have reported that longer or more frequent naps were linked to a higher risk of dementia and faster cognitive decline,^5,47^ while others have found a lower risk^48,49^ or no association.^7,50^ Our study demonstrated that long and frequent napping, when combined with good nighttime sleep dimensions, might not affect the risk of dementia. This underscores the importance of clustering analysis and considering combination of sleep and circadian dimensions, as longer and more frequent naps alone were associated with a higher risk of dementia in our sample. Furthermore, this is in line with a previous clustering study which showed that a “high sleep propensity” group (characterized by long naps) was protective against all-cause mortality, while napping alone was associated with a higher risk.^28^ Interestingly, LFN were linked to increased risk of CVD events, although the association was of marginal significance. Prior research on napping and CVD has been mixed, with several studies suggesting a higher risk of CVD associated with more frequent or longer naps,^4,12^ while others suggested a protective effect.^13^ Daytime napping may result from short or poor nighttime sleep (as a compensatory mechanism) or indicate poor overall health, both of which can contribute to increase CVD risk. However, these hypotheses do not fully explain our findings since LFN had normal nighttime sleep duration with good sleep quality, and LFN did not differ from AHS regarding sociodemographic factors and comorbidities. Although the exact reasons why LFN might be associated with CVD but not dementia are not well-understood, assumptions include autonomic nervous system disruptions or other metabolic changes not examined in this study,^51,52^ which may impact more the cardiovascular risk. It may also involve cardiovascular mechanisms that do not relate to dementia risk or have a less direct effect on it. Further research, including mediation analyses, is needed to better understand the role of napping in relation to adverse health outcomes and their underlying mechanisms.

Our findings have important clinical and public health implications. By identifying common multidimensional sleep and circadian patterns in older men using advanced machine learning techniques, this study enhances our understanding of the interrelations between numerous sleep/circadian parameters and underscores the critical need for comprehensive sleep health assessment in clinical practice and research settings. Both FPS and LFN exhibited poor circadian activity rhythmicity, emphasizing the importance of this dimension of sleep health. Future studies should incorporate circadian rhythms when examining adverse outcomes. Moreover, our results highlight specific at-risk groups that could benefit from sleep interventions and prevention efforts, and support poor sleep patterns as a marker or risk factor for cognitive and cardiovascular health. Public health initiatives may consider prioritizing the screening and monitoring of older adults with weak circadian rhythms combined with poor nighttime sleep or with high daytime napping.

## Strengths and Limitations

Strengths of this study include a 12-year longitudinal design with high retention rates, a multidimensional measure of sleep and rest-activity rhythms using objective measures, and consideration of numerous potential confounders. We also used an innovative machine learning approach capable of detecting clusters with flexible shapes, which standard clustering methods cannot achieve. However, there are also limitations. The diagnosis of dementia relied on cognitive tests and self-reported data, which may lead to outcome misclassification. Moreover, the timing of dementia incidence was based on study visit dates, which may not reflect the actual onset of dementia, and information on dementia subtypes was lacking. This study predominantly involves White older men, limiting the generalizability of the results. Future research should replicate these methods in more diverse samples. Lastly, as an observational study, we cannot assume causal relationships between sleep profiles and dementia or CVD events.

## Conclusions

In older men, we identified three multidimensional actigraphy-derived sleep/circadian profiles. Compared to AHS, FPS were associated with less favorable cognitive and cardiovascular health over 12 years, while FPS were linked to increased risk of CVD events, but not dementia. These results suggest potential targets for sleep interventions and prevention efforts and emphasize the need for careful screening of poor sleepers for adverse outcomes. Moreover, our study highlights the importance of future research to consider combinations of sleep characteristics.

## Data Availability

The data supporting the findings of this study are openly available at https://mrosonline.ucsf.edu.

## Acknowledgment

The authors thank the study staff and all the men who participated in MrOS Sleep Study.

## Data Sharing

The data supporting the findings of this study are openly available at https://mrosonline.ucsf.edu.

