## Supplementary Materials for "Multidimensional Sleep Profiles via Machine learning and Risk of Dementia and Cardiovascular Disease"

eMethods

**Additional Information about Cardiovascular Disease Event**

For fatal events, the death certificate and hospital records from the time of death were collected. For fatal events that were not hospitalized, a proxy interview with next of kin and hospital records from the most recent hospitalization in the last 12 months were obtained. These documents were used to determine the underlying cause of death.

Confirmed cardiovascular events were grouped as follows: 1) Coronary heart disease event: acute myocardial infarction (ST or non-ST elevation), sudden coronary heart disease death, coronary artery bypass surgery, mechanical coronary revascularization, hospitalization for unstable angina, ischemic congestive heart failure, or other coronary heart disease event not described above; 2) Cerebrovascular event: stroke or transient ischemic attack; 3) Peripheral vascular disease event: acute arterial occlusion, rupture, dissection, or vascular surgery; and 4) All-cause cardiovascular disease event: combines coronary heart disease, cerebrovascular event, and peripheral vascular disease.

**Covariate Data**

All participants completed questionnaires at baseline, which included items about demographics, smoking status, caffeine intake (mg/day), and alcohol use (>1 alcoholic drink/week). Level of physical activity was assessed using the Physical Activity Scale for the Elderly.^1^ Participants also self-reported their medical history, specifically prior physician diagnosis of heart attack, stroke, diabetes mellitus, and hypertension. The Geriatric Depression Scale was used to assess depressive symptoms, with higher scores corresponding to higher levels of depression.^2^ Participants were asked to bring in all medications used within the preceding 30 days. All prescription and nonprescription medications were entered into an electronic database and each medication was matched to its ingredient(s) based on the Iowa Drug Information Service Drug Vocabulary (College of Pharmacy, University of Iowa, Iowa City, IA).^3^ The use of antidepressants, benzodiazepines, and other sleep medications (non-benzodiazepines, non-barbiturate sedative hypnotics) were categorized. A comprehensive examination included measurements of body weight and height. Body mass index was calculated as weight in kilograms divided by the square of height in meters.

**Additional Information about Statistical Analysis**

*Sleep Profiles Identification*

We conducted a cluster analysis to identify distinct sleep/circadian profiles. Firstly, we selected variables for inclusion in the analysis, considering the high correlation among the actigraphy variables (eFigure 2). When the correlation coefficient between two variables was >0.70, we retained only one of the variables prioritized on clinical meaningfulness, resulting in a final selection of 20 sleep/circadian variables. Secondly, we performed a principal component analysis (PCA) on the selected variables to reduce data dimensionality (while preserving most of the data variation) and enhance the efficacy of subsequent clustering. The number of principal components (PCs) was determined according to the Kaiser-Guttman criteria,^4^ considering PCs with eigenvalues >1, and complemented by visual inspection of the scree plot to identify the “elbow” point in the eigenvalues distribution (eFigure 3).^5^ Thirdly, sleep/circadian profiles were identified using Multiple Coalesced Generalized Hyperbolic Distribution (MCGHD; MixGHD package in R) mixture models based on the six PCs derived from the PCA.^6,7^ This method, as opposed to standard clustering approaches, was chosen for its ability to accommodate potentially skewed and/or asymmetric clusters, an important consideration given the skewed distributions often observed in actigraphy data (see eFigure 4 for distributions). We explored models comprising one to five clusters, using k-medoids as the starting criterion, and determined the optimal number of clusters by examining the Bayesian Information Criteria (BIC), the Akaike Information Criteria (AIC) and the Integrated Complete-data Likelihood (ICL) (higher values indicating a better fit for the data). The optimal number of clusters was determined by selecting solutions displaying an “elbow” in the AIC and BIC plots and/or a subsequent drop in ICL.

Sleep and circadian characteristics were compared across clusters using the Kruskal-Wallis test. We calculated effect sizes using the eta-squared (η^2^) statistic to characterize the magnitude of cluster differences. The effect was considered small for η^2^ <0.06, moderate for 0.06 ≤ η^2^ <0.14, and large for η^2^ ≥0.14.^8^ In post-hoc analyses, we performed multiple pairwise-comparisons using Dunn’s test adjusted for multiple comparisons with the Bonferroni correction.

*Time to Dementia and CVD events*

We assessed the proportional hazard assumption for each independent variable by examining the Schoenfeld residuals. If a variable violated the assumption, we carried out a stratified Cox regression model for that specific variable.

**eTable 1**. Baseline sample characteristics between included and excluded men.

|  | **Included**  **(n= 2,667)** | **Excluded**  **(n=468)** |  |
| --- | --- | --- | --- |
| **Characteristics** | **Median (IQR) or No. (%)** | **Median (IQR) or No. (%)** | **p-value^a^** |
| Age (years) | 75 (72;80) | 77 (73;82) | <.0001 |
| Education, *≤High school* | 539 (20.2) | 127 (27.1) | 0.0007 |
| Race/ethnicity |  |  | <.0001 |
| *White* | 2425 (90.9) | 391 (83.5) |  |
| *Black/African American* | 82 (3.1) | 39 (8.3) |  |
| *Other* | 160 (6.0) | 38 (8.1) |  |
| PASE score | 142.5 (97.2;187.2) | 130.6 (88.5;184.1) | 0.01 |
| GDS score, *≥6* | 156 (5.9) | 55 (11.8) | <.0001 |
| Smoking status |  |  | 0.46 |
| *Never* | 1054 (39.5) | 181 (38.8) |  |
| *Past* | 1561 (58.6) | 273 (58.5) |  |
| *Current* | 51 (1.9) | 13 (2.8) |  |
| Caffeine intake (mg/day) | 184 (36;368) | 144 (0;314) | 0.045 |
| Alcoholic drink per week, *>1* | 1452 (54.7) | 203 (43.8) | <.0001 |
| BMI | 26.8 (24.6;29.4) | 26.2 (24.3;29.4) | 0.18 |
| History of heart attack | 450 (16.9) | 90 (19.3) | 0.21 |
| History of stroke | 93 (3.5) | 24 (5.1) | 0.08 |
| History of diabetes mellitus | 352 (13.2) | 65 (13.9) | 0.67 |
| History of hypertension | 1324 (49.7) | 236 (50.5) | 0.73 |
| Current sleep medication, *≥1* | 304 (11.4) | 89 (19.1) | <.0001 |
| Antidepressants | 187 (7.0) | 61 (13.1) | <.0001 |
| BZD | 107 (4.0) | 32 (6.9) | 0.006 |
| Other sleep medications | 50 (1.9) | 12 (2.6) | 0.32 |

Abbreviations: BZD, benzodiazepine; BMI, body mass index; GDS, Geriatric Depression Scale; IQR, interquartile range; PASE, Physical Activity Scale for the Elderly.

^a^ Kruskal-Wallis test was used for continuous variables, Chi-square test for categorical variables

**eTable 2**. Cox regression models of the association between sleep profiles and the incidence of dementia (n=2,562) and cardiovascular disease events (n=2,606).

|  | Unadjusted model | |  | Model 1 | |  | Model 2 | |
| --- | --- | --- | --- | --- | --- | --- | --- | --- |
|  | HR (95% CI) | p |  | HR (95% CI) | p |  | HR (95% CI) | p |
| Dementia |  | 0.09 |  |  | 0.11 |  |  | 0.08 |
| Active healthy sleepers | 1 |  |  | 1 |  |  | 1 |  |
| Fragmented poor sleepers | 1.34 (1.03;1.74) | 0.03 |  | 1.35 (1.02;1.78) | 0.03 |  | 1.39 (1.04;1.85) | 0.02 |
| Frequent and long nappers | 1.11 (0.89;1.39) | 0.37 |  | 1.09 (0.86;1.38) | 0.48 |  | 1.11 (0.87;1.42) | 0.41 |
| Cardiovascular disease events |  | 0.0004 |  |  | 0.01 |  |  | 0.007 |
| Active healthy sleepers | 1 |  |  | 1 |  |  | 1 |  |
| Fragmented poor sleepers | 1.44 (1.19;1.74) | 0.0002 |  | 1.32 (1.08;1.60) | 0.006 |  | 1.34 (1.10;1.63) | 0.003 |
| Frequent and long nappers | 1.21 (1.02;1.42) | 0.02 |  | 1.16 (0.98;1.37) | 0.08 |  | 1.18 (1.00;1.39) | 0.05 |

Age was used as time scale. Model 1 was adjusted for site, race/ethnicity, education, smoking status, caffeine intake, alcohol use, physical activity, body mass index, history of diabetes mellitus and hypertension, depressive symptoms, and sleep-related medications use. Model 2 was further adjusted for history of heart attack and stroke.

**eTable 3**. Cox regression models of the association between sleep profiles and the incidence of dementia (n=2,424) and cardiovascular disease events (n=2,455) after adjusting for baseline apnea-hypopnea index (AHI).

|  | Model 1 | |  | Model 2 | |
| --- | --- | --- | --- | --- | --- |
|  | HR (95% CI) | p |  | HR (95% CI) | p |
| Dementia |  | 0.15 |  |  | 0.11 |
| Active healthy sleepers | 1 |  |  | 1 |  |
| Fragmented poor sleepers | 1.33 (1.00;1.78) | 0.05 |  | 1.38 (1.02;1.86) | 0.04 |
| Frequent and long nappers | 1.11 (0.87;1.41) | 0.41 |  | 1.14 (0.88;1.47) | 0.31 |
| Cardiovascular disease events |  | 0.006 |  |  | 0.003 |
| Active healthy sleepers | 1 |  |  | 1 |  |
| Fragmented poor sleepers | 1.38 (1.13;1.69) | 0.001 |  | 1.41 (1.15;1.72) | 0.0008 |
| Frequent and long nappers | 1.15 (0.97;1.37) | 0.11 |  | 1.17 (0.99;1.40) | 0.07 |

Age was used as time scale. Model 1 was adjusted for site, race/ethnicity, education, smoking status, caffeine intake, alcohol use, physical activity, body mass index, history of diabetes mellitus and hypertension, depressive symptoms, sleep-related medications use, and baseline AHI. Model 2 was further adjusted for history of heart attack and stroke.

**eTable 4**. Cox regression models of the association between sleep profiles and dementia incidence after adjusting for baseline Modified Mini-Mental State Examination (3MS) score (n=2,562).

|  | Model 1 | |  | Model 2 | |
| --- | --- | --- | --- | --- | --- |
|  | HR (95% CI) | p |  | HR (95% CI) | p |
| Dementia |  | 0.16 |  |  | 0.11 |
| Active healthy sleepers | 1 |  |  | 1 |  |
| Fragmented poor sleepers | 1.32 (1.00;1.74) | 0.049 |  | 1.37 (1.03;1.82) | 0.03 |
| Frequent and long nappers | 1.07 (0.84;1.35) | 0.58 |  | 1.09 (0.85;1.40) | 0.49 |

Age was used as time scale. Model 1 was adjusted for site, race/ethnicity, education, smoking status, caffeine intake, alcohol use, physical activity, body mass index, history of diabetes mellitus and hypertension, depressive symptoms, sleep-related medications use, and baseline 3MS score. Model 2 was further adjusted for history of heart attack and stroke.

**eTable 5**. Cox regression models of the association between sleep profiles and dementia incidence after exclusion of incident dementia cases identified at the first follow-up visit (n=2,403).

|  | Unadjusted model | |  | Model 1 | |
| --- | --- | --- | --- | --- | --- |
|  | HR (95% CI) | p |  | HR (95% CI) | p |
| Dementia |  | 0.10 |  |  | 0.10 |
| Active healthy sleepers | 1 |  |  | 1 |  |
| Fragmented poor sleepers | 1.41 (1.03;1.95) | 0.03 |  | 1.42 (1.01;1.98) | 0.04 |
| Frequent and long nappers | 1.17 (0.89;1.54) | 0.26 |  | 1.20 (0.90;1.61) | 0.20 |

Age was used as time scale. Model 1 was adjusted for site, race/ethnicity, education, smoking status, caffeine intake, alcohol use, physical activity, body mass index, history of diabetes mellitus and hypertension, depressive symptoms, and sleep-related medications use.

**eTable 6**. Cox regression models of the association between sleep profiles and the incidence of cardiovascular disease events after exclusion of participants with a history of heart attack or stroke (n=2,106).

|  | Unadjusted model | |  | Model 1 | |
| --- | --- | --- | --- | --- | --- |
|  | HR (95% CI) | p |  | HR (95% CI) | p |
| Cardiovascular disease events |  | 0.003 |  |  | 0.04 |
| Active healthy sleepers | 1 |  |  | 1 |  |
| Fragmented poor sleepers | 1.48 (1.19;1.85) | 0.0004 |  | 1.36 (1.08;1.70) | 0.009 |
| Frequent and long nappers | 1.12 (0.91;1.36) | 0.28 |  | 1.07 (0.87;1.31) | 0.51 |

Age was used as time scale. Model 1 was adjusted for site, race/ethnicity, education, smoking status, caffeine intake, alcohol use, physical activity, body mass index, history of diabetes mellitus and hypertension, depressive symptoms, and sleep-related medications use. Model 2 was further adjusted for history of heart attack and stroke.

**eFigure 1**. Flow chart.

Men enrolled in MrOS Study

N = **5,994**

Men enrolled in ancillary Sleep Visit 1

N = **3,135**

**2,859** not at the Sleep Visit 1:

1,997 Refused

349 Died

40 Terminated

150 Ineligible

323 Not asked. Recruitment goal met.

n Not Determined:

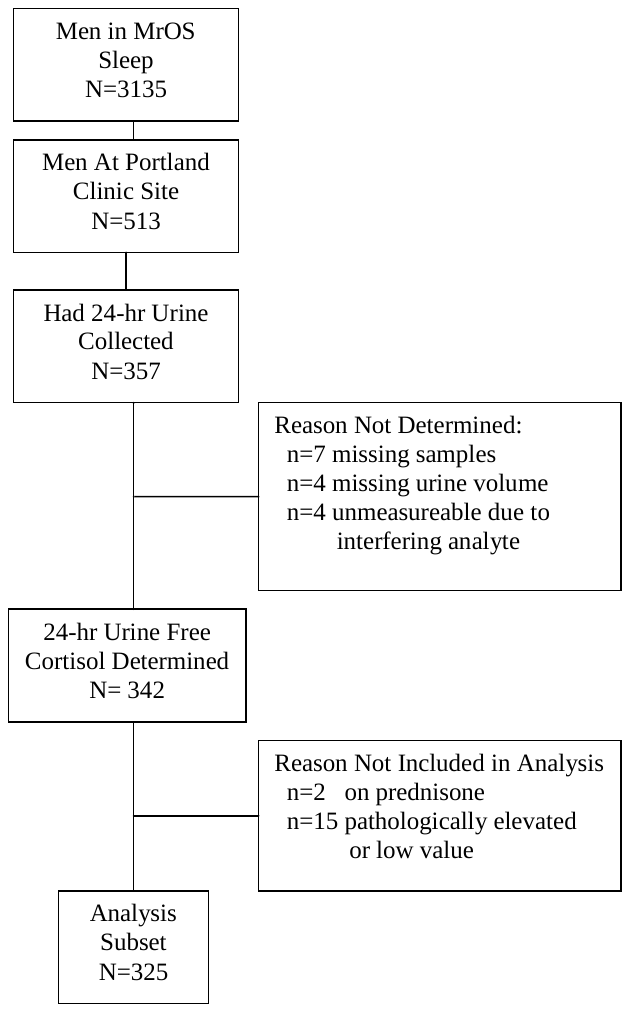

n=7 missing samples

n=4 missing urine volume

n=4 unmeasureable due to

interfering analyte

**468** Missing data/excluded at Sleep Visit 1:

331 invalid actigraphy data

294 with ≤ 3 valid “in-bed” and “out-of-bed” intervals

37 missing actigraphy data

137 excluded for probable dementia (use of dementia medication or 3MS test score < 80)

Cognitively intact men at Sleep Visit 1 with actigraphy data

N = **2,667**

**eFigure 2**. Heatmap.

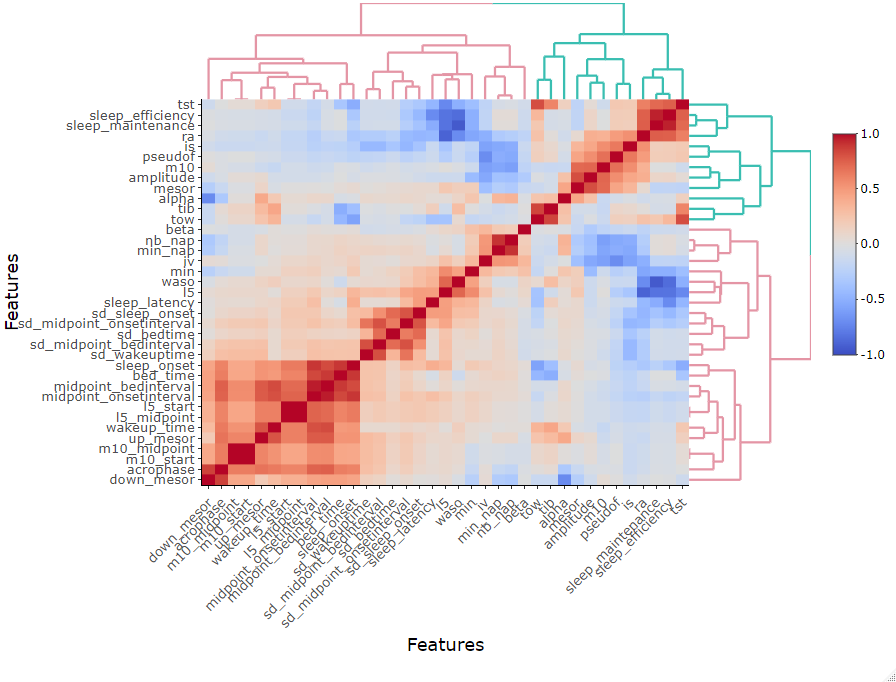

**eFigure 3**. Criteria for determining the number of principal components.

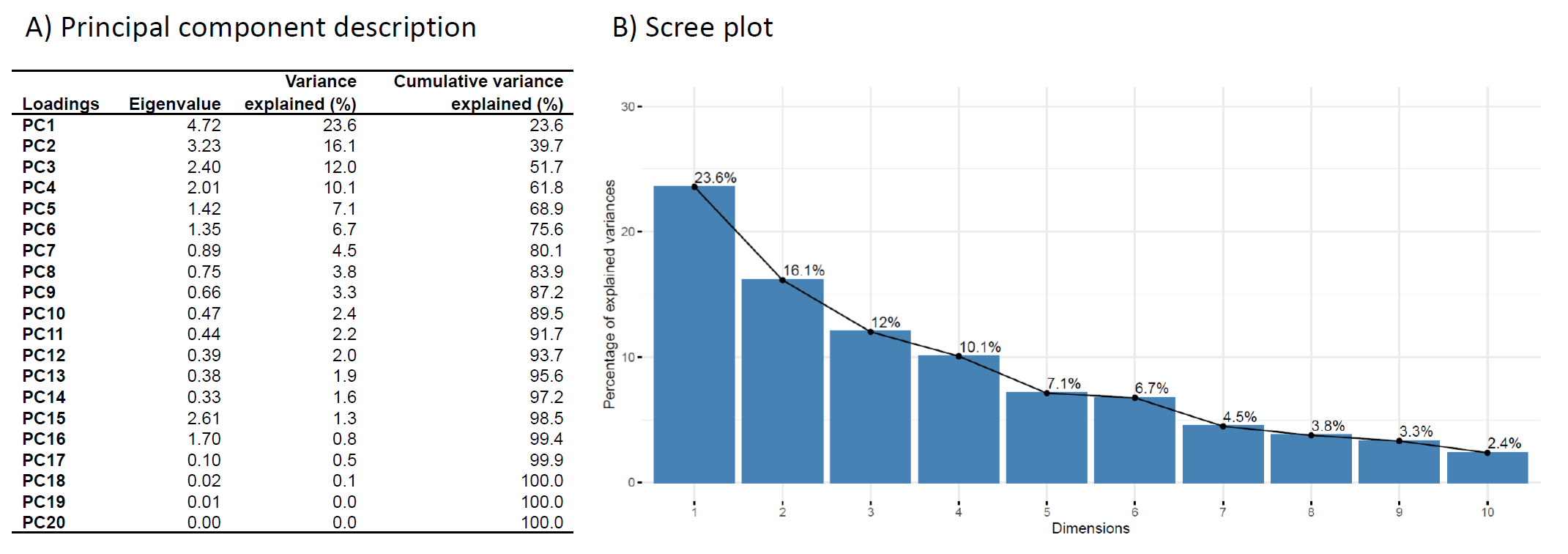

Abbreviations: PC, principal component.

**eFigure 4**. Distributions of actigraphy variables used to identify the sleep clusters.

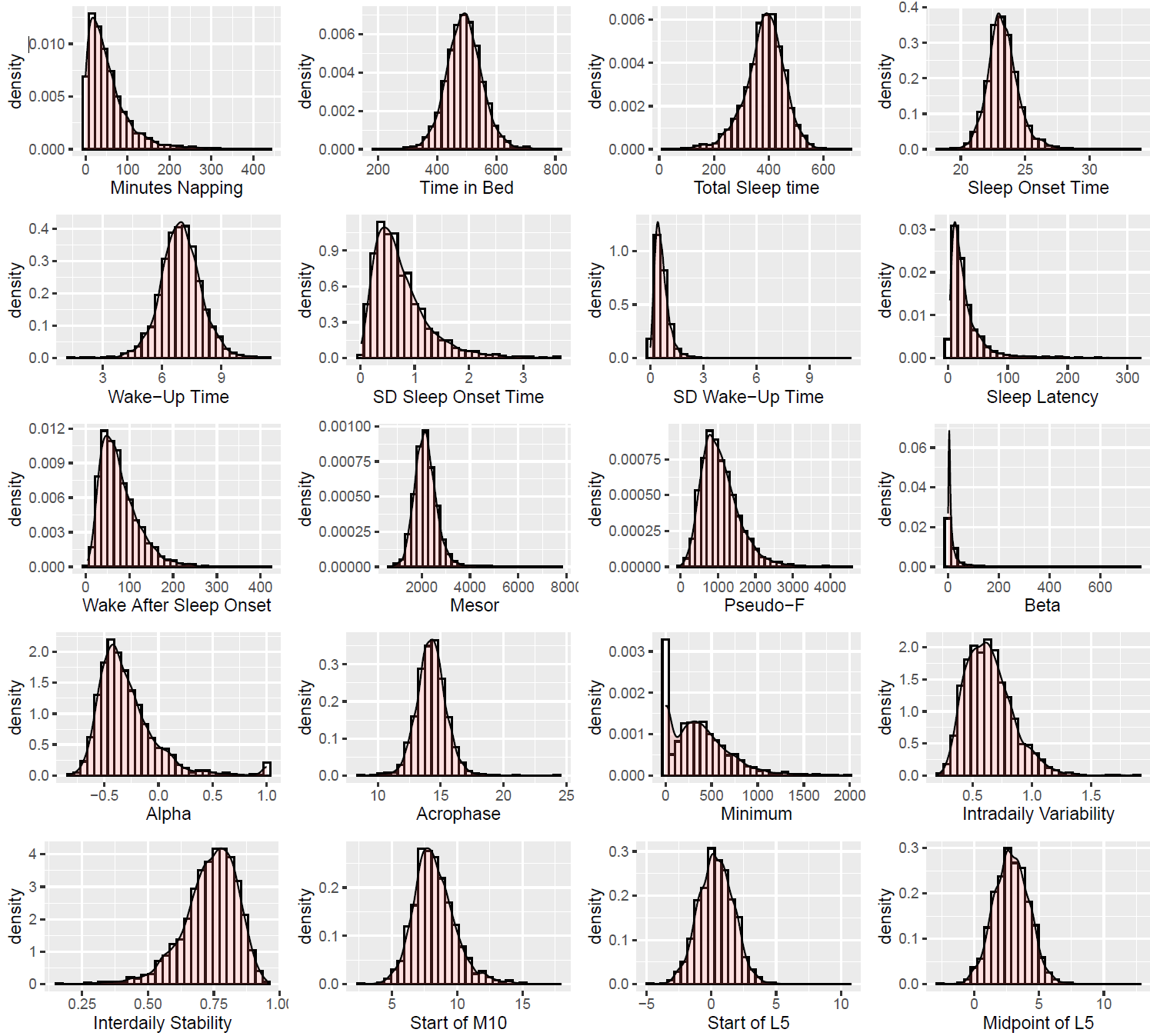

**eFigure 5**. Criteria for determining the number of clusters.

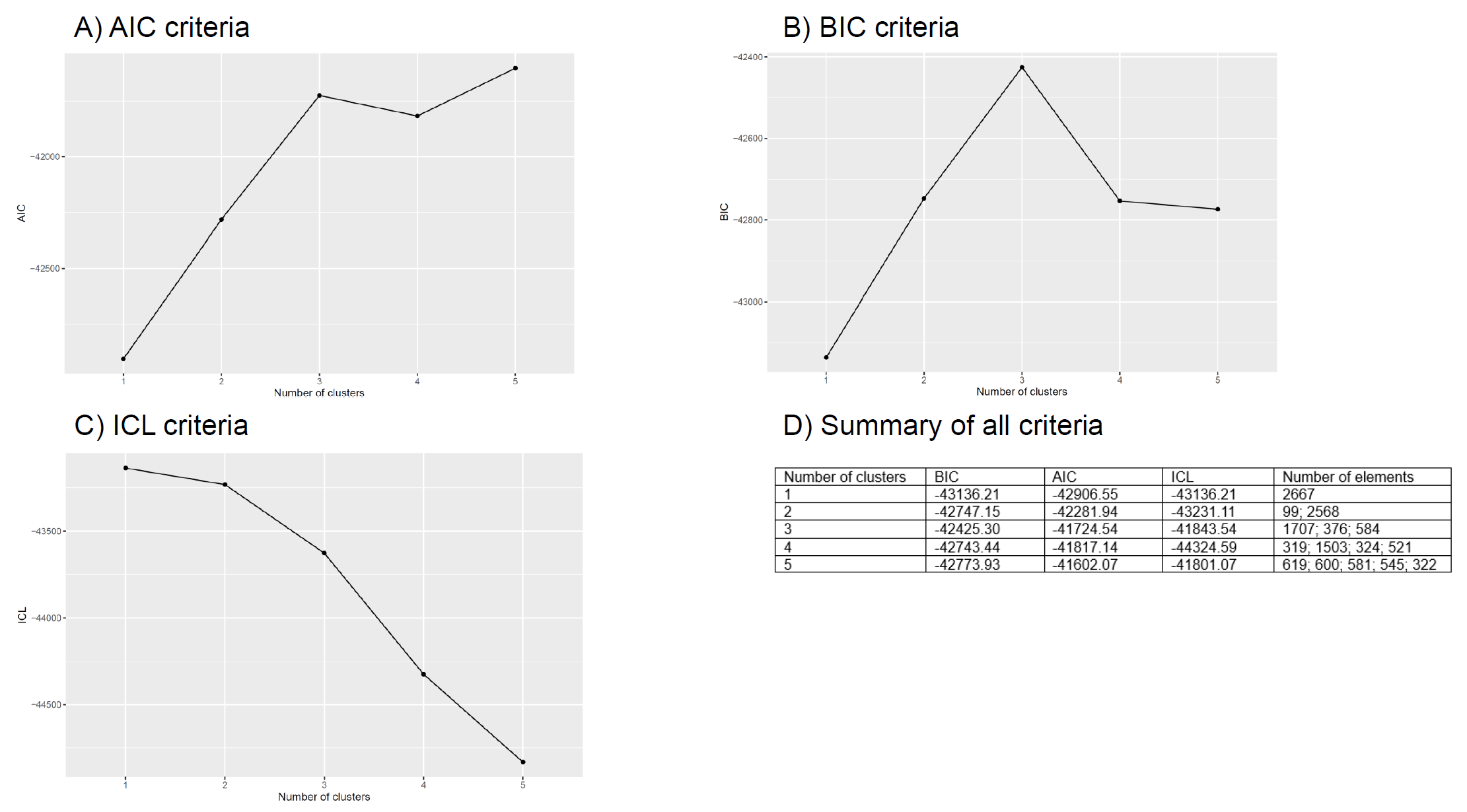

Abbreviations: AIC, Akaike Information Criteria; BIC, Bayesian Information Criteria; ICL, Integrated Complete-data Likelihood.
